# Pre-stroke glucagon-like peptide-1 receptor agonist use and outcomes after acute ischemic stroke: a propensity score-matched retrospective cohort study

**DOI:** 10.64898/2026.04.29.26352109

**Authors:** Yasmin Negida, Yugant Khand, Yousef Hawas, Vidit Yadav, Khaled M.H. Mohamed, Ram Saha

**Affiliations:** Virginia Commonwealth University School of Medicine, Richmond, Virginia, USA; Department of Neurology, Mayo Clinic Florida, Jacksonville, Florida, USA; Faculty of Medicine, Tanta University, Tanta, Egypt; Department of Internal Medicine, Mayo Clinic Florida, Jacksonville, Florida, USA; College of Health and Human Sciences, North Dakota State University, Fargo, North Dakota, USA

**Author notes:** Corresponding author: Yasmin Negida, MD | Department of Neurology, Virginia Commonwealth University, Richmond, VA, USA | |.

**Keywords:** GLP-1 receptor agonist, acute ischemic stroke, type 2 diabetes mellitus, mortality, intracerebral hemorrhage, neuroprotection, propensity score matching

## Abstract

**Background:** Glucagon-like peptide-1 receptor agonists (GLP-1 RAs) have demonstrated cardiovascular benefits in type 2 diabetes mellitus (T2DM); however, their association with post-stroke outcomes following acute ischemic stroke (AIS) remains uncertain.

**Methods:** We conducted a retrospective propensity score-matched (PSM) cohort study using the TriNetX Global Collaborative Network. Adults with T2DM and AIS between January 2020 and January 2025 were included. Pre-stroke GLP-1 RA users were compared with non-users. Primary outcomes were all-cause mortality, intracerebral hemorrhage (ICH), ICD-10-coded stroke severity (NIHSS categories), and transient ischemic attack (TIA).

**Results:** After PSM, GLP-1 RA use was associated with significantly lower all-cause mortality at all time points (RR 0.44–0.52; HR 0.43–0.45; all p<0.001). ICH risk was reduced within 1–3 days (RR 0.683; 95% CI 0.496–0.940; p=0.019) and within 7 days (RR 0.575; 95% CI 0.516–0.641; p<0.001). A severity-dependent gradient was observed across NIHSS categories, with risk reductions ranging from 37% for minor strokes (NIHSS 0–5; RR 0.626) to 67% for very severe strokes (NIHSS >20; RR 0.327; all p<0.001). TIA risk was 29% lower (RR 0.712; 95% CI 0.668–0.759; p<0.001). E-value analysis demonstrated that an unmeasured confounder would need to be associated with both GLP-1 RA use and 30-day mortality by a risk ratio of at least 3.95 to fully explain the observed association, and by a risk ratio of at least 3.53 to shift the confidence interval to include the null.

**Conclusions:** In this large real-world cohort, pre-stroke GLP-1 RA use was associated with lower post-stroke mortality, reduced ICH, and a severity-dependent reduction in ICD-10-coded stroke severity among patients with T2DM. Residual confounding cannot be excluded, and these findings warrant confirmation in prospective randomized trials.

## Introduction

Stroke remains the second leading cause of death and the third leading contributor to long-term disability globally, accounting for approximately 7.3 million deaths and 160.5 million disability-adjusted life-years annually [1]. Despite advances in acute stroke management and secondary prevention, effective neuroprotective strategies remain a critical unmet need [2].

Type 2 diabetes mellitus (T2DM) is associated with worse functional outcomes, higher mortality, and greater rates of hemorrhagic transformation following AIS [3,4]. Prospective meta-analyses report a 2-to 2.3-fold increase in stroke incidence among individuals with T2DM compared with those without [5]. Mechanistically, chronic hyperglycemia induces endothelial dysfunction, oxidative stress, prothrombotic clot-architecture alterations, and impaired collateral circulation [6,7].

Glucagon-like peptide-1 receptor agonists (GLP-1 RA), widely used in the management of type 2 diabetes mellitus, have demonstrated cardiovascular benefits extending beyond glycemic control. They demonstrated significant reductions in major adverse cardiovascular events (MACE), with a subsequent reduction in MACE and reduction in stroke risk [8–10]. GLP-1 receptors are expressed on cerebral endothelial cells and neurons; their activation enhances cerebral blood flow, reduces infarct volume, attenuates neuroinflammation via microglial modulation, and protects the blood-brain barrier against hemorrhagic transformation in experimental stroke models [11–13]. Furthermore, exendin-4, a GLP-1 RA, has been shown to reduce hemorrhagic transformation in both thrombolytic and anticoagulant models of experimental stroke [14].

Despite this mechanistic rationale, large-scale real-world evidence evaluating GLP-1 RA use in the context of acute stroke outcomes is lacking. Whether pre-stroke GLP-1 RA exposure modifies post-stroke mortality, hemorrhagic complications, or stroke severity in unselected patients with T2DM remains unknown. We therefore conducted a large retrospective propensity score-matched cohort study using the TriNetX Global Collaborative Network to examine these associations between pre-stroke GLP-1 RA use and key post-stroke outcomes in patients with T2DM.

## Methods

### Study Design and Data Source

This was a retrospective PSM cohort study conducted using the TriNetX Global Collaborative Network, a federated research platform aggregating de-identified electronic health record (EHR) data from over 100 healthcare organizations across multiple countries. Data extraction and analysis were performed within the TriNetX platform between February 2026 and March 2026. The study was reported in accordance with the Strengthening the Reporting of Observational Studies in Epidemiology (STROBE) guidelines [15]. The TriNetX platform operates under a business associate agreement with participating institutions. The study was exempt from institutional review board approval as confirmed by the Virginia Commonwealth University IRB.

### Study Populatio

We identified adults aged ≥18 years with a documented diagnosis of T2DM (ICD-10-CM E11) who subsequently developed AIS (ICD-10-CM I63) between January 1, 2020, and January 1, 2025. Counts are reported as approximate owing to TriNetX platform rounding policies. The total study population comprised approximately 474,000 patients. The exposure cohort included approximately 31,000 patients with documented GLP-1 RA use prior to the index AIS event; the control cohort comprised approximately 443,000 patients with T2DM and AIS and no documented GLP-1 RA exposure.

### Exposure Definition

GLP-1 RA exposure was defined as any documented prescription or administration of a GLP-1 RA (including liraglutide, semaglutide, dulaglutide, exenatide, lixisenatide, or tirzepatide [a dual GIP/GLP-1 receptor agonist]) recorded in the EHR prior to the index AIS event. To minimize immortal time bias, cohort entry was aligned at the date of AIS diagnosis for both groups, with exposure status determined from pre-stroke records. Exposure timing relative to stroke (e.g., recent versus remote use), treatment duration, and adherence could not be reliably ascertained within the TriNetX platform. Agent-level analyses were not performed due to the aggregate nature of exposure coding within the TriNetX platform.

### Outcomes

Time zero (index date) was defined as the date of AIS diagnosis. Patients were followed from the index date until outcome occurrence or censoring at prespecified time points. Primary outcomes were: (1) all-cause mortality at same-day, 30-day, 60-day, and 90-day time points; (2) intracerebral hemorrhage (ICH; ICD-10-CM I61) within 1–3 days and within 7 days of index AIS, serving as a surrogate for hemorrhagic transformation; (3) ICD-10-coded stroke severity using the NIHSS R29.7-series codes across five categories: minor (NIHSS 0–5), mild (6–10), moderate (11–15), severe (16–20), and very severe (>20); and (4) TIA (ICD-10-CM G45). An exploratory analysis of recurrent cerebral infarction (ICD-10-CM I63) within five years was conducted but was deemed unreliable owing to probable capture of index event codes (see Results). Competing risks were not explicitly modeled; accordingly, non-fatal outcome estimates may be influenced by differential mortality between groups, with the direction of bias likely attenuating effect sizes for non-fatal endpoints. Given 13 primary comparisons, the possibility of type I error inflation should be considered when interpreting results.

### Propensity Score Matching

To address confounding by indication, 1:1 nearest-neighbor PSM with a caliper width of 0.1 standard deviations was performed separately for each outcome to maximize statistical power and maintain covariate balance across endpoints. Covariates included age at index event, sex, race, ethnicity, hypertension, heart failure, atrial fibrillation, ischemic heart disease, body mass index (BMI), glycated hemoglobin (HbA1c), statin therapy, anticoagulant therapy, and tobacco exposure. Covariate balance was assessed using standardized mean differences (SMDs); adequate balance was defined as all SMDs <0.1 after matching.

### Statistical Analysis

Between-group comparisons were performed using measures of association including risk ratios (RR) with 95% confidence intervals (CI) and absolute risk differences (ARD). Time-to-event analyses were conducted using Kaplan-Meier survival estimation with hazard ratios (HR) derived from Cox proportional hazards models, and between-group differences assessed by the log-rank test. All analyses were performed within the TriNetX analytics platform. Statistical significance was defined as a two-sided p-value <0.05. Missing data were handled using complete case analysis as per TriNetX platform constraints.

### E-Value Analysis

To quantify the robustness of primary findings to potential unmeasured confounding, E-value analysis was performed for the 30-day mortality outcome (the most clinically consequential primary endpoint) using the risk ratio scale. E-values were calculated for both the point estimate and the confidence interval limit closest to the null, following the method of VanderWeele and Ding. An E-value represents the minimum strength of association (on the risk ratio scale) that an unmeasured confounder would need to have with both GLP-1 RA use and mortality to fully explain the observed association.

### Sensitivity Analysis: Exclusion of Thrombolysis Recipients

We performed sensitivity analysis to account for potential confounding from reperfusion therapy by excluding patients who received intravenous thrombolysis (alteplase or tenecteplase). Cohorts were redefined to exclude these agents at baseline, and the analysis was repeated using identical inclusion criteria, outcome definitions, and propensity score matching methodology. ICH within 7 days was re-evaluated using both risk-based and time-to-event approaches to assess the robustness of the primary findings independent of thrombolytic exposure.

### NIHSS Coding Considerations

NIHSS categories were ascertained from ICD-10-CM diagnostic codes within the TriNetX platform. Patients with a relevant NIHSS code documented prior to the analysis time window were excluded from the respective analysis. NIHSS documentation rates in administrative data are low (0.3%–7.6% across severity categories in this cohort), reflecting incomplete coding in EHRs. Accordingly, NIHSS severity outcomes represent the presence of a coded NIHSS category rather than a systematically recorded clinical score, and analyses were restricted to patients with available coding. No imputation for missing NIHSS data was performed [16].

## Results

### Study Population and Baseline Characteristics

A total of approximately 474,000 adults with T2DM and AIS were identified, of whom approximately 31,000 (6.5%) had documented pre-stroke GLP-1 RA use. Before propensity score matching (PSM), GLP-1 RA users were significantly younger (mean age 67.5 vs. 73.0 years; SMD 0.47), and had higher prevalence of hypertension (87.2% vs. 68.8%; SMD 0.45), ischaemic heart disease (45.2% vs. 35.2%; SMD 0.21), heart failure (26.8% vs. 20.8%; SMD 0.14), and greater statin use (87.5% vs. 54.7%; SMD 0.77). They also had higher mean HbA1c (8.16% vs. 7.14%; SMD 0.51) and BMI (33.4 vs. 30.0 kg/m^2^; SMD 0.46). These differences reflect a population with a greater burden of cardiometabolic disease among GLP-1 RA users, consistent with channeling bias toward higher-risk patients. After outcome-specific PSM, all covariates achieved adequate balance (maximum SMD <0.1 across 14 variables), indicating successful mitigation of baseline differences (Table 1).

**Table 1.** Outcome-Specific Propensity Score-Matched Cohort Sizes.

| Outcome | Time Window | GLP-1 RA (n) | Control (n) | Total Matched |
| --- | --- | --- | --- | --- |
| Mortality | Same day | 29,448 | 29,429 | 58,877 |
| Mortality | 30-day | 29,922 | 29,909 | 59,831 |
| Mortality | 60-day | 29,922 | 29,909 | 59,831 |
| Mortality | 90-day | 29,640 | 29,640 | 59,280 |
| ICH | 1–3 days | 28,328 | 28,234 | 56,562 |
| ICH | 7 days | 28,747 | 28,970 | 57,717 |
| NIHSS 0–5 | Post-stroke | 29,207 | 29,207 | 58,414 |
| NIHSS 6–10 | Post-stroke | 29,495 | 29,842 | 59,337 |
| NIHSS 11–15 | Post-stroke | 29,892 | 30,004 | 59,896 |
| NIHSS 16–20 | Post-stroke | 29,069 | 29,129 | 58,198 |
| NIHSS >20 | Post-stroke | 29,105 | 29,118 | 58,223 |
| TIA | Post-stroke | 30,147 | 30,147 | 60,294 |
| Recurrent CI† | 5 years | 27,190 | 27,190 | 54,380 |

†Recurrent cerebral infarction analysis deemed unreliable; see Results. CI, cerebral infarction; ICH, intracerebral hemorrhage; NIHSS, National Institutes of Health Stroke Scale; TIA, transient ischemic attack.

### All-Cause Mortality

Pre-stroke GLP-1 RA use was associated with a 55% relative reduction in mortality across all time points. Same-day mortality occurred in 31 of 29,448 GLP-1 RA users (0.11%) compared with 60 of 29,429 controls (0.20%), yielding a risk ratio of 0.516 (95% CI 0.335–0.796; p=0.002). The Kaplan-Meier hazard ratio was not estimated at this time point owing to insufficient events for stable survival curve estimation.

At 30 days, mortality occurred in 596 of 29,922 GLP-1 RA users (1.99%) versus 1,347 of 29,909 controls (4.50%), corresponding to a risk ratio of 0.442 (95% CI 0.402–0.486; p<0.001) and a hazard ratio of 0.433 (95% CI 0.393–0.477; log-rank p<0.001). The magnitude of effect was consistent at 60 days (RR 0.461, 95% CI 0.425–0.500; HR 0.448, 95% CI 0.413–0.487) and 90 days (RR 0.452, 95% CI 0.419–0.488; HR 0.437, 95% CI 0.404–0.472; all p<0.001; Figure 2).

The absolute risk reduction at 90 days was 3.69%, corresponding to a number needed to treat (NNT) of 27 (Table 2).

**Table 2.**
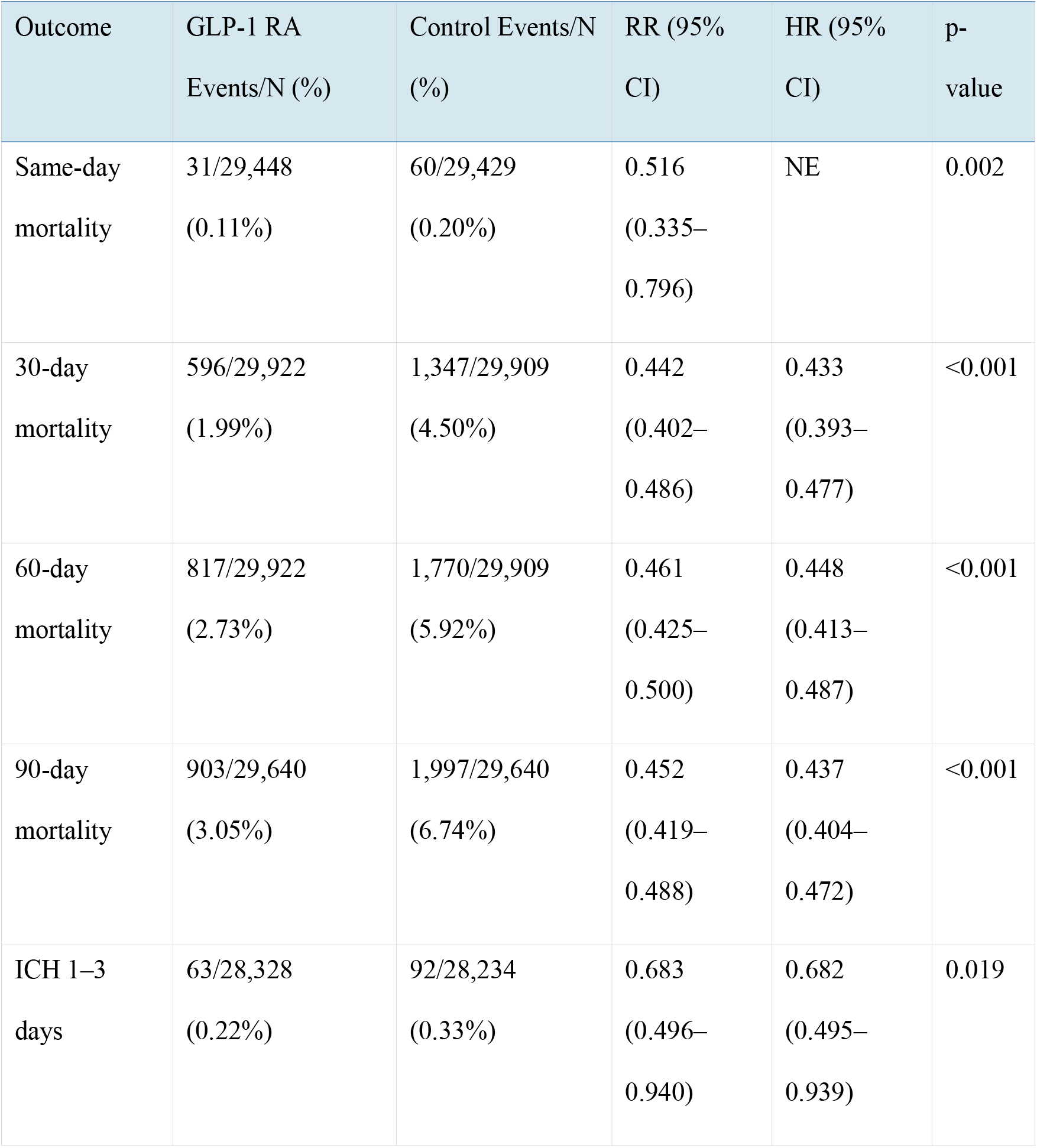
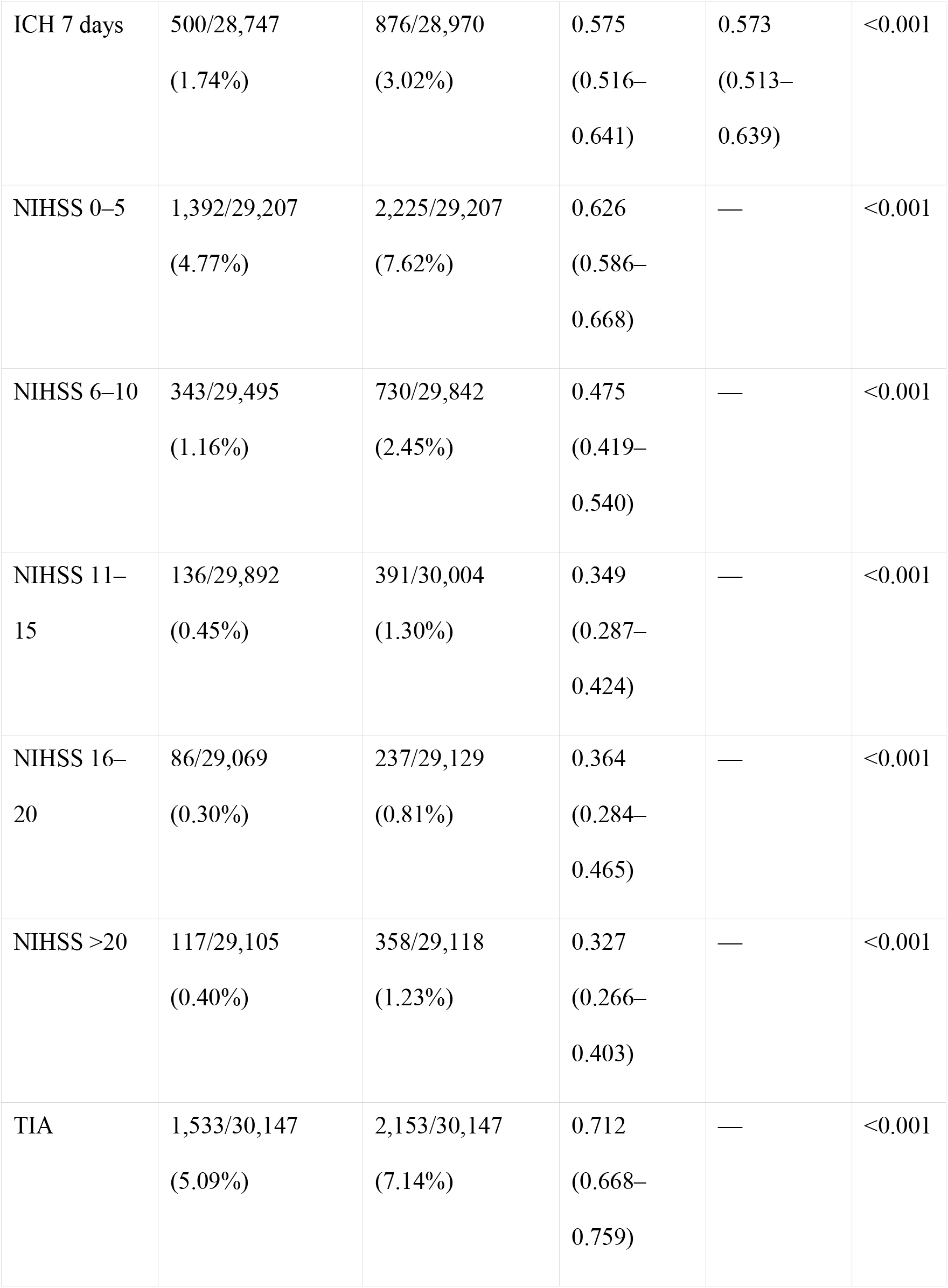
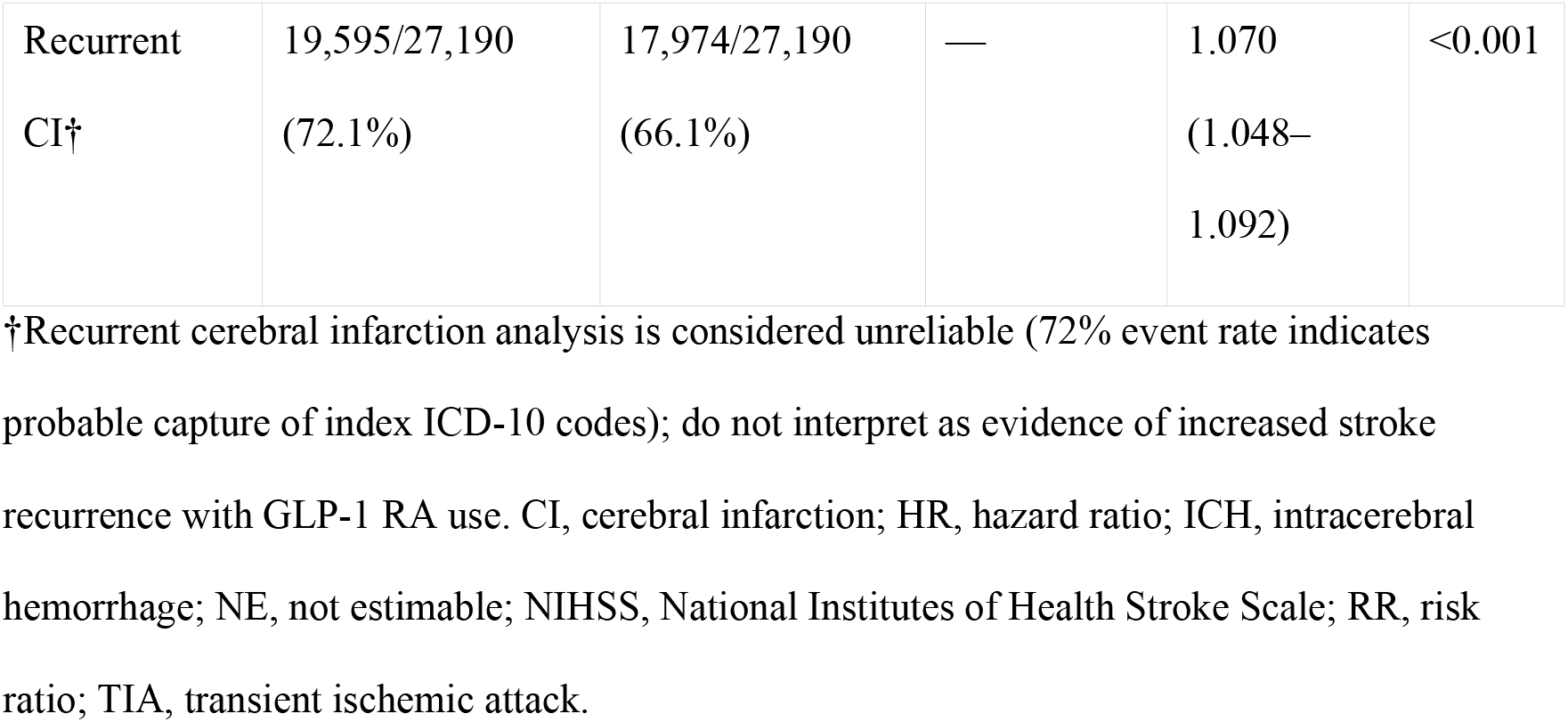
Summary of Primary and Exploratory Outcomes After Propensity Score Matching.

| Outcome | GLP-1 RA<br>Events/N (%) | Control Events/N<br>(%) | RR (95%<br>CI) | HR (95%<br>CI) | p-value |
| --- | --- | --- | --- | --- | --- |
| Same-day<br>mortality | 31/29,448<br>(0.11%) | 60/29,429<br>(0.20%) | 0.516<br>(0.335–<br>0.796) | NE | 0.002 |
| 30-day<br>mortality | 596/29,922<br>(1.99%) | 1,347/29,909<br>(4.50%) | 0.442<br>(0.402–<br>0.486) | 0.433<br>(0.393–<br>0.477) | <0.001 |
| 60-day<br>mortality | 817/29,922<br>(2.73%) | 1,770/29,909<br>(5.92%) | 0.461<br>(0.425–<br>0.500) | 0.448<br>(0.413–<br>0.487) | <0.001 |
| 90-day<br>mortality | 903/29,640<br>(3.05%) | 1,997/29,640<br>(6.74%) | 0.452<br>(0.419–<br>0.488) | 0.437<br>(0.404–<br>0.472) | <0.001 |
| ICH 1–3<br>days | 63/28,328<br>(0.22%) | 92/28,234<br>(0.33%) | 0.683<br>(0.496–<br>0.940) | 0.682<br>(0.495–<br>0.939) | 0.019 |
| ICH 7 days | 500/28,747<br>(1.74%) | 876/28,970<br>(3.02%) | 0.575<br>(0.516–<br>0.641) | 0.573<br>(0.513–<br>0.639) | <0.001 |
| NIHSS 0–5 | 1,392/29,207<br>(4.77%) | 2,225/29,207<br>(7.62%) | 0.626<br>(0.586–<br>0.668) | — | <0.001 |
| NIHSS 6–10 | 343/29,495<br>(1.16%) | 730/29,842<br>(2.45%) | 0.475<br>(0.419–<br>0.540) | — | <0.001 |
| NIHSS 11–<br>15 | 136/29,892<br>(0.45%) | 391/30,004<br>(1.30%) | 0.349<br>(0.287–<br>0.424) | — | <0.001 |
| NIHSS 16–<br>20 | 86/29,069<br>(0.30%) | 237/29,129<br>(0.81%) | 0.364<br>(0.284–<br>0.465) | — | <0.001 |
| NIHSS >20 | 117/29,105<br>(0.40%) | 358/29,118<br>(1.23%) | 0.327<br>(0.266–<br>0.403) | — | <0.001 |
| TIA | 1,533/30,147<br>(5.09%) | 2,153/30,147<br>(7.14%) | 0.712<br>(0.668–<br>0.759) | — | <0.001 |
| Recurrent<br>CI† | 19,595/27,190<br>(72.1%) | 17,974/27,190<br>(66.1%) | — | 1.070<br>(1.048–<br>1.092) | <0.001 |

†Recurrent cerebral infarction analysis is considered unreliable (72% event rate indicates probable capture of index ICD-10 codes); do not interpret as evidence of increased stroke recurrence with GLP-1 RA use. CI, cerebral infarction; HR, hazard ratio; ICH, intracerebral hemorrhage; NE, not estimable; NIHSS, National Institutes of Health Stroke Scale; RR, risk ratio; TIA, transient ischemic attack.

E-value analysis for the 30-day mortality finding demonstrated that an unmeasured confounder would need to be associated with both GLP-1 RA use and mortality by a risk ratio of at least 3.95 to fully explain the observed association (RR 0.442), and by a risk ratio of at least 3.53 to shift the confidence interval to include the null. These thresholds exceed the effect sizes of most measured confounders in similar datasets, supporting the robustness of this finding to residual confounding, although they do not exclude the possibility of strong unmeasured bias.

### Intracerebral Hemorrhage (ICH)

In the primary analysis, GLP-1 RA use was associated with a significantly lower risk of ICH following acute ischemic stroke. Within 1–3 days, ICH occurred in 63 of 28,328 GLP-1 RA users (0.22%) compared with 92 of 28,234 non-users (0.33%), with a reduced risk ratio of 0.683, (95% CI 0.496–0.940; p=0.019) and hazard ratio of 0.682, 95% (CI 0.495–0.939; log-rank p=0.018). ICH incidence within 7 days remained lower among GLP-1 RA users (1.74% vs 3.02%; RR 0.58, 95% CI 0.52–0.64; HR 0.57, 95% CI 0.51–0.64; p<0.001). Sensitivity analyses excluding patients receiving intravenous thrombolysis showed consistent risk reduction, with a 0.6% absolute risk difference favoring GLP-1 RA use (Figure 1).

**Figure 1.**
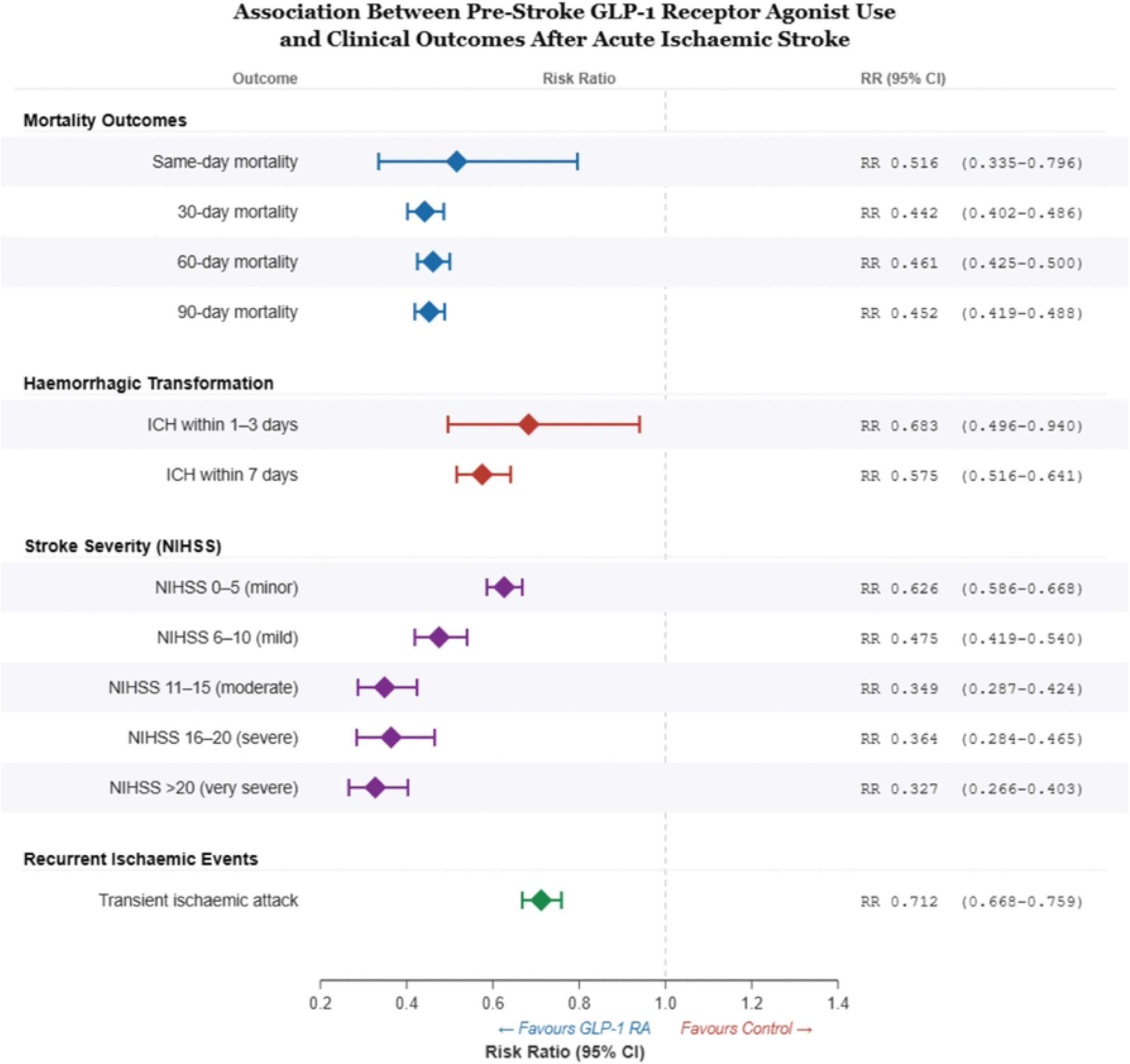
Forest plot of risk ratios for all primary outcomes. Risk ratios with 95% confidence intervals are shown for all-cause mortality (same-day, 30-day, 60-day, 90-day), intracerebral hemorrhage (1–3 days, 7 days), ICD-10-coded stroke severity by NIHSS category (five categories), and transient ischemic attack in propensity score-matched cohorts. All outcomes show risk ratios <1.0, favoring GLP-1 RA users. The dashed vertical line indicates RR = 1.0. Point estimates are displayed as filled squares; horizontal lines indicate 95% confidence intervals.

## NIHSS-Coded Stroke Severity

There was a consistent and severity-dependent gradient observed across all five NIHSS categories. For minor strokes (NIHSS 0–5), 1,392 of 29,207 GLP-1 RA users (4.77%) had the outcome code documented compared with 2,225 of 29,207 controls (7.62%; RR 0.626, 95% CI 0.586–0.668; p<0.001). The risk ratio decreased progressively with increasing severity: mild strokes (NIHSS 6–10; RR 0.475, 95% CI 0.419–0.540), moderate strokes (NIHSS 11–15; RR 0.349, 95% CI 0.287–0.424), severe strokes (NIHSS 16–20; RR 0.364, 95% CI 0.284–0.465), and very severe strokes (NIHSS >20; RR 0.327, 95% CI 0.266–0.403; all p<0.001; Figure 1). “Time-to-event analysis was not applicable for NIHSS categories, as these represent categorical documentation rather than longitudinal outcomes. Documentation rates ranged from 0.30% (severe, NIHSS 16–20) to 7.62% (minor, NIHSS 0–5 in controls). Although these low ascertainment rates limit the absolute prevalence estimates, the consistent direction and graded magnitude of the associations across all five categories strengthen confidence in the findings.

### Transient Ischemic Attack

TIA was documented in 1,533 of 30,147 GLP-1 RA users (5.09%) compared with 2,153 of 30,147 controls (7.14%), yielding a risk ratio of 0.712 (95% CI 0.668–0.759; p<0.001; Figure 1). This finding was consistent in direction with other outcomes, supporting a broader cerebrovascular protective association.

### Recurrent Cerebral Infarction (Exploratory)

The exploratory analysis of recurrent stroke yielded implausibly high event rates (>70%), likely reflecting misclassification of index events. This outcome was therefore deemed unreliable and is not interpreted further. In matched cohorts (n=27,190 per group), the event rate was 72.1% (19,595/27,190) in GLP-1 RA users versus 66.1% (17,974/27,190) in controls, with a hazard ratio of 1.070 (95% CI 1.048–1.092; log-rank p<0.001). The implausibly high event rates in both groups strongly suggest that the ICD-10 I63 codes captured the index stroke event itself rather than truly new recurrent events, as no exclusion of patients with prior ICD-10 I63 codes was applied in this analysis (unlike the other outcome analyses). The modest HR >1 favouring controls is therefore likely an artifact of differential coding patterns. This outcome should not be interpreted as evidence that suggests GLP-1 RAs increase stroke recurrence.

## Discussion

In 474,000 patients with T2DM and AIS, pre-stroke GLP-1 RA use was associated with significantly lower post-stroke mortality (48–56% relative risk reduction), reduced intracerebral haemorrhage (32–43% reduction), a severity-dependent gradient in NIHSS-coded stroke severity (37–67% reduction), and lower rates of TIA (29% reduction). These associations were consistent across analysis and persisted after PSM for a broad range of clinical covariates.

The mortality reductions observed in our study (RR 0.44–0.52) are substantially larger than the 12% MACE reduction and 16% stroke reduction reported in the Kristensen et al. meta-analysis of seven cardiovascular outcome trials [10]. Several factors may account for this discrepancy. This could be since the trial-based estimates reflect chronic cardiovascular protection in populations selected for high cardiovascular risk but not specifically for acute cerebrovascular events. In contrast, our study examines outcomes in patients who have already experienced AIS.

In such cases, there is acute neuroprotection owing to reduction of excitotoxicity, neuroinflammation, and blood-brain barrier disruption which may contribute to additional benefit beyond chronic atherosclerotic risk reduction [11,12,17,18]. Furthermore, the larger effect sizes may partially reflect healthy user bias, whereby patients receiving GLP-1 RAs may be more engaged with the healthcare system and have better overall disease management, despite our efforts to adjust for measured confounders. GLP-1 RA users may differ in unmeasured ways, including healthcare engagement, socioeconomic status, medication adherence, and access to acute stroke care. These factors could contribute to improved outcomes independent of pharmacologic effects.

The reduction in ICH risk among GLP-1 RA users represents, to our knowledge, a novel clinical finding. While previous cardiovascular outcome trials reported neutral effects on haemorrhagic stroke, the present analysis specifically examined ICH occurring within days of AIS. This timeframe is consistent with haemorrhagic transformation of the index infarct rather than primary haemorrhagic stroke. However, detailed data on acute reperfusion therapies were limited, despite their well-established role in influencing stroke outcomes. To address this, we conducted a sensitivity analysis excluding thrombolysis-treated patients. The persistence of a similar effect size for intracerebral hemorrhage suggests that the observed association is unlikely to be explained by differences in reperfusion therapy alone. The 32% reduction at 1–3 days (RR 0.683) and 43% reduction at 7 days (RR 0.575) align with preclinical evidence demonstrating that GLP-1 RAs protect the blood-brain barrier and reduce haemorrhagic transformation. Liu et al. showed that exendin-4 reduced haemorrhagic transformation in a thrombolytic model of experimental stroke [14], while Chen et al. demonstrated similar protection in a warfarin-associated haemorrhagic transformation model [19]. These preclinical findings provide biological plausibility for the clinical observation and suggest that GLP-1 RA-mediated blood-brain barrier protection may translate to reduced haemorrhagic complications in the acute post-stroke period. Xie et al. further demonstrated the role of CD147-mediated pathways in haemorrhagic transformation, providing additional mechanistic context [20]. Although consistent with preclinical data, the observational nature of this finding precludes causal inference.

Perhaps the most intriguing finding is the severity-dependent gradient across NIHSS categories, with risk reductions increasing from 37% for minor strokes (NIHSS 0–5) to 67% for very severe strokes (NIHSS >20). This gradient has important implications for interpretation. While this pattern is biologically plausible, it should be interpreted cautiously given low NIHSS coding rates and the potential for differential documentation. This pattern is consistent with preclinical observations that GLP-1 RAs reduce infarct volume and neuronal death in a dose-dependent manner [18,21], and that GLP-1 receptor activation modulates microglial phenotype from pro-inflammatory to anti-inflammatory states, an effect that may be more consequential in larger infarcts with greater neuroinflammatory burden [11].

However, several caveats temper this interpretation. NIHSS documentation rates were low (0.3–7.6%), indicating that these findings apply only to the subset of patients with coded NIHSS data and may not be representative of the broader population. Furthermore, differential coding practices between GLP-1 RA users and non-users cannot be excluded, although there is no established mechanism by which GLP-1 RA prescription status would influence NIHSS documentation behaviour.

The observed effects are consistent with multiple mechanistic pathways. GLP-1 receptors are expressed on cerebral endothelial cells and neurons, and their activation has been shown to enhance cerebral blood flow through nitric oxide-dependent vasodilation of cerebral arterioles[13]. Activation of neuronal GLP-1 receptors promotes anti-apoptotic signalling and reduces excitotoxic neuronal death [12,22]. Moreover, GLP-1 RAs have been demonstrated to modulate microglial polarization, reducing pro-inflammatory cytokine release and promoting a neuroprotective microenvironment [18]. Also, proteomic analyses have identified GLP-1-regulated pathways involved in synaptic plasticity and neuronal survival [23]. Additionally, dual GLP-1/GIP receptor agonists have shown complementary neuroprotective effects in experimental stroke models [24]. GLP-1 RAs were found to mitigate stroke-related hyperglycaemia and the associated oxidative stress, both of which are established predictors of poor stroke outcomes [25].

### 5.1 Clinical Implications

These findings generate a strong hypothesis that GLP-1 RAs may function as disease-modifying agents in AIS, rather than solely preventive therapies. Given the widespread and increasing use of GLP-1 RA globally, even modest effects on post-stroke outcomes could have substantial public health implications. However, changes to prescribing practice cannot be recommended based on observational data alone. Additionally, the findings remained significant after excluding thrombolysis treated patients, indicating that the observed association is not explained by differences in reperfusion therapy.

### 5.2 Strengths, Limitations, and Future Directions

To our knowledge, this study represents the largest real-world analysis of GLP-1 RA use and post-stroke outcomes, with approximately 474,000 patients. The outcome-specific PSM approach maximized sample size for each analysis while ensuring covariate balance (all SMDs <0.1). The examination of multiple outcomes (mortality, ICH, NIHSS severity, TIA) with consistent direction of effect strengthens the overall conclusion. The novel findings regarding ICH reduction and the severity-dependent NIHSS gradient provide new clinical insights and generate testable hypotheses for prospective investigation. These findings suggest that GLP-1 RA therapy may confer benefits extending beyond chronic risk reduction, potentially influencing acute stroke trajectories. Randomized controlled trials specifically designed to evaluate peri-stroke GLP-1 RA exposure are urgently needed.

Several important limitations must be acknowledged. First, this is a retrospective observational study, and despite rigorous PSM, residual confounding from unmeasured variables cannot be excluded. Healthy user biases in which GLP-1 RA users may represent a more health-conscious population and have greater access to healthcare, remains an important concern. Second, the study relies on ICD-10-CM codes, which are subject to coding variability and may not capture the full clinical picture. NIHSS coding in administrative databases is notably incomplete, with documentation rates as low as 0.3% in some categories. Third, we lacked data on functional outcomes (e.g., modified Rankin Scale), stroke aetiology (e.g., TOAST classification), and acute stroke treatments (e.g., intravenous thrombolysis, mechanical thrombectomy), all of which may influence outcomes and could confound our results. Fourth, the study period (2020–2025) overlaps with the COVID-19 pandemic, which may have influenced stroke presentations, hospital care patterns, and mortality. Fifth, we could not determine the specific GLP-1 RA agent, dose, duration of use, or adherence, precluding dose-response or agent-specific analyses. Finally, the recurrent cerebral infarction analysis was compromised due to probable capture of index event codes, highlighting the limitations of administrative data for this particular outcome.

## Conclusions

In this large propensity score-matched cohort study of patients with T2DM and AIS, pre-stroke GLP-1 RA use was associated with substantially lower post-stroke mortality, reduced intracerebral haemorrhage, and a severity-dependent reduction in NIHSS-coded stroke severity. The magnitude of these associations appears larger than expected based on cardiovascular risk reduction alone, although residual confounding cannot be excluded. With over 30 million patients worldwide currently prescribed GLP-1 RAs, these findings have significant clinical implications and warrant confirmation in prospective randomized trials specifically designed to evaluate the neuroprotective potential of GLP-1 RAs in AIS.

## Data Availability

The data that support the findings of this study are available from the TriNetX Global Collaborative Network. Restrictions apply to the availability of these data, which were used under license for this study and are not publicly available. Data are, however, available from TriNetX (https://trinetx.com) to qualified researchers with appropriate access.

## Acknowledgments

The authors acknowledge the TriNetX Global Collaborative Network and its participating healthcare organizations for providing access to the de-identified electronic health record data used in this study.

## Data Availability

The data that support the findings of this study are available from the TriNetX Global Collaborative Network (https://trinetx.com) under license. These data are not publicly available; however, they are accessible to qualified researchers through institutional access. No individual-level data can be shared due to platform restrictions.

## Author Contributions

Yasmin Negida: Conceptualization, study design, data analysis, interpretation, manuscript drafting. Yugant Khand: Methodology, data interpretation, critical revision. Yousef Hawas: Data curation, manuscript drafting, critical revision. Vidit Yadav: Methodology, manuscript drafting, critical revision. Khaled M.H. Mohamed: Literature review, manuscript drafting, critical revision. Ram Saha: Supervision, study oversight, critical revision.

## Ethics Approval

The Virginia Commonwealth University Institutional Review Board determined that this study does not constitute human subjects research due to the use of de-identified data from the TriNetX platform; therefore, institutional review board approval and informed consent were not required.

